# A Noncooperative Game Analysis for Controlling COVID-19 Outbreak

**DOI:** 10.1101/2020.05.22.20110783

**Authors:** Anupam Kumar Bairagi, Mehedi Masud, Do Hyeon Kim, Md. Shirajum Munir, Abdullah Al Nahid, Sarder Fakhrul Abedin, Kazi Masudul Alam, Sujit Biswas, Sultan S Alshamrani, Zhu Han, Choong Seon Hong

## Abstract

*COVID-19* is a global epidemic. Till now, there is no remedy for this epidemic. However, isolation and social distancing are seemed to be effective to control this pandemic. In this paper, we provide an analytical model on the effectiveness of the sustainable lockdown policy that accommodates both isolation and social distancing features of the individuals. To promote social distancing, we analyze a noncooperative game environment that provides an incentive for maintaining social distancing. Furthermore, the sustainability of the lockdown policy is also interpreted with the help of a game-theoretic incentive model for maintaining social distancing. Finally, an extensive numerical analysis is provided to study the impact of maintaining a social-distancing measure to prevent the Covid-19 outbreak. Numerical results show that the individual incentive increases more than 85% with an increasing percentage of home isolation from 25% to 100% for all considered scenarios. The numerical results also demonstrate that in a particular percentage of home isolation, the individual incentive decreases with an increasing number of individuals.

## 1 Introduction

The novel Coronavirus (2019-nCoV or COVID-19) is one of the most dangerous pandemics of this century. COVID-19 has already affected every aspect of individual’s life i.e. politics, sovereignty, economy, education, religion, entertainment, sports, tourism, transportation, and manufacturing. It was first identified in Wuhan City, China on December 29, 2019, and after a short span of time, it broke out worldwide [1,2]. The World Health Organization (WHO) has announced the COVID-19 outbreak as a Public Health Emergency of International Concern (PHEIC) and identified it as an epidemic on January 30, 2020 [3]. COVID-19 has affected 210 countries and territories throughout the globe and 2 international conveyances [4,6] till April 20, 2020.

COVID-19 was first exposed to Wuhan, China massively, and, then, it spread to other areas of China, and later to other parts of the planet. Until 20 April 2020, COVID-19 has affected more than 2, 415, 370 persons [3–6] and has become the most critical global affair. Meanwhile, the total number of death and recovery to/from COVID-19 are 165, 903 and 632, 484, respectively till April 20, 2020 [3–6]. Different countries are undertaking different initiatives to fight against the COVID-19 epidemic, but there is no clear-cut solution to date. Among the worldwide recovery of 632, 484 humans from COVID-19, only China reported 77,062 recovery cases [3–6]. So far, the recovery cases against the infected cases are not satisfactory globally anyway.

One of the most crucial tasks that countries need to do for understanding and preventing the spread of COVID-19 is testing. Testing allows infected bodies to acknowledge that they are already affected. This can be helpful for taking care of them, and also to decrease the possibility of contaminating others. Testing is also essential for a proper reply to the pandemic. It allows carrying evidence-based steps to slow down the spread of COVID-19. However, to date, the testing capability for COVID-19 is quite inadequate in most countries around the world. South Korea was the second COVID-19 infectious country after China during February 2020. However, mass testing may be one of the reasons why it succeeded to diminish the number of new infections since it facilitates a rapid identification of potential outbreaks [8]. For detecting COVID-19, two kinds of tests are clinically carried out: (i) detection of virus particles in swabs collected from the mouth or nose and (ii) estimates the antibody response to the virus in blood serum.

This COVID-19 epidemic is still uncontrolled in most countries. Infected cases and death graph are rising every day. However, researchers are also focusing on the learning-based mechanism for detecting COVID-19 infections [16–22]. This approach can be cost-effective and also possibly will take less time to perform the test. Other studies [9–14] focus on analyzing the epidemiological and/or clinical characteristics of COVID-19. As per our knowledge, there is no study that focuses on mathematical model is for monitoring and controlling individual to prevent this COVID-19 epidemic. Thus, the main contribution of this paper is to develop an effective mathematical model with the help of global positioning system (GPS) information to fight against COVID-19 epidemic by monitoring and controlling individual. To this end, we make the following key contributions:

- First, we study the real-world dataset to realize the worldwide severity of COVID-19 epidemic and also show the predicted results for infected and active cases of COVID-19.
- Second, we consider an optimization problem that maximizes the social utility of individuals by taking into account isolation and social distancing policies. Here, the optimization parameters are the positions of an individual.
- Third, we analyze the objective function by incorporating the social distancing feature of an individual in a noncooperative game environment. Here, we observe that home isolation is the dominant strategy for all the players of the game.
- Finally, we interpret the sustainability of lockdown policy for controlling the impact of COVID-19 outbreak through extensive numerical analysis.

The remainder of the paper is organized as follows. The global phenomenon of COVID-19 is presented in Section 2. In Section 3, we present the literature review. We explain the system model and present the problem formulation in Section 4. The consider solution approach of the above-mentioned problem is addressed in Section 5. We interpret the sustainability of lockdown policy with our model in Section 6. In Section 7, we provide an extensive numerical analysis to study the impact of maintaining a social-distancing measure. Finally, we draw the conclusions in Section 8.

## 2 Analytical Study on Global Phenomenon of COVID-19

In this subsection, we present the analysis of available COVID-19 data [3–6]. Fig. 1 shows that the daily cumulative infected, death, recovered, and active cases of all over the world till April 17, 2020, and showing the significant increase of confirmed cases over time. In fact, a sharp increase in infected cases from the third week of March 2020 is observed in Fig. 1. This is due to the massive spread of COVID-19 incumbents in Europe and USA. Fig. 2 shows the percentage of top 10 infected countries of COVID-19 till April 18, 2020, and they contributed 77% of inmates over all the infected cases.

**Fig 1.**
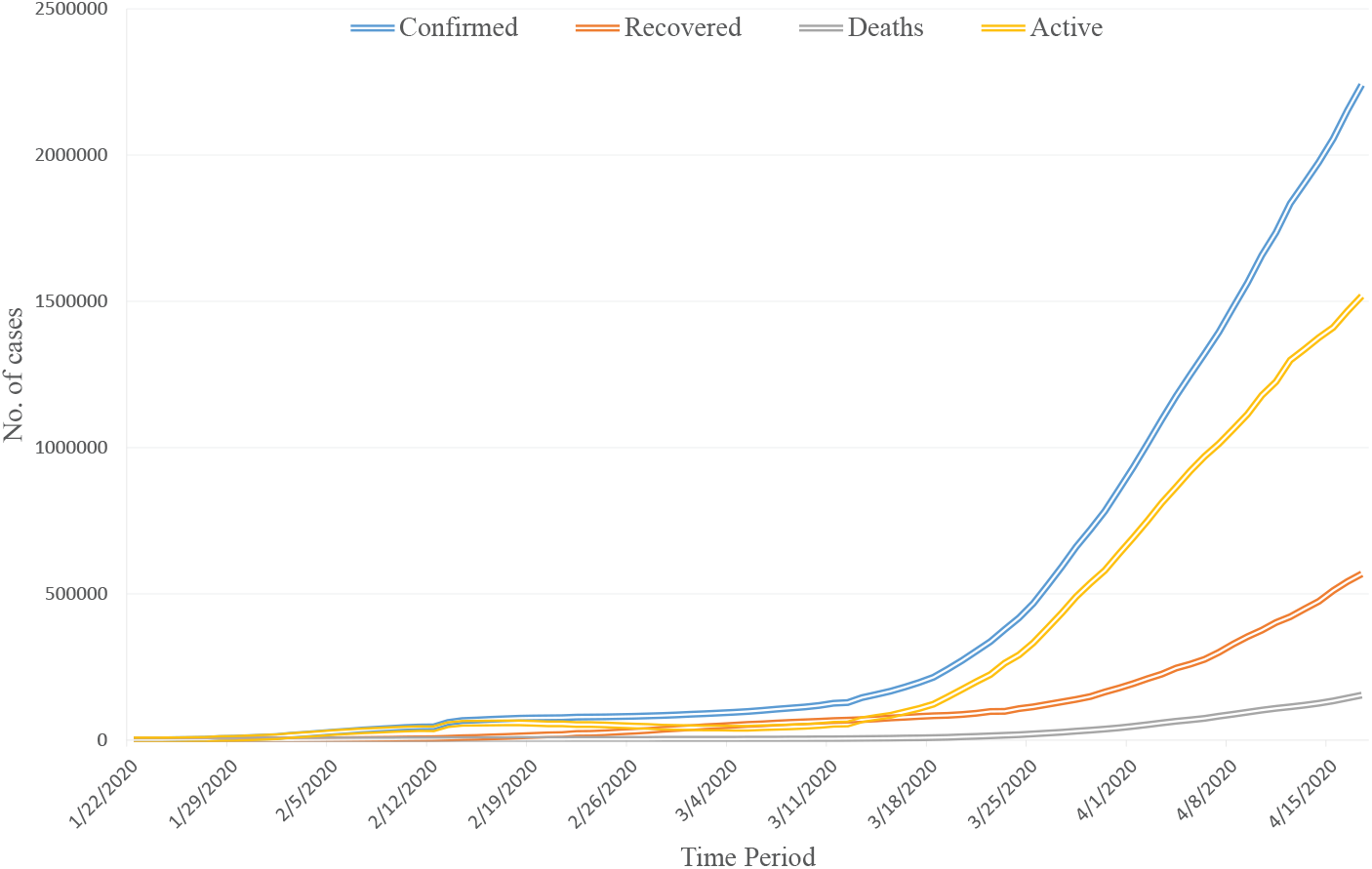
Daily cumulative confirmed, death, recovered, and active cases of COVID-19 from January 22, 2020 to April 17, 2020.

**Fig 2.**
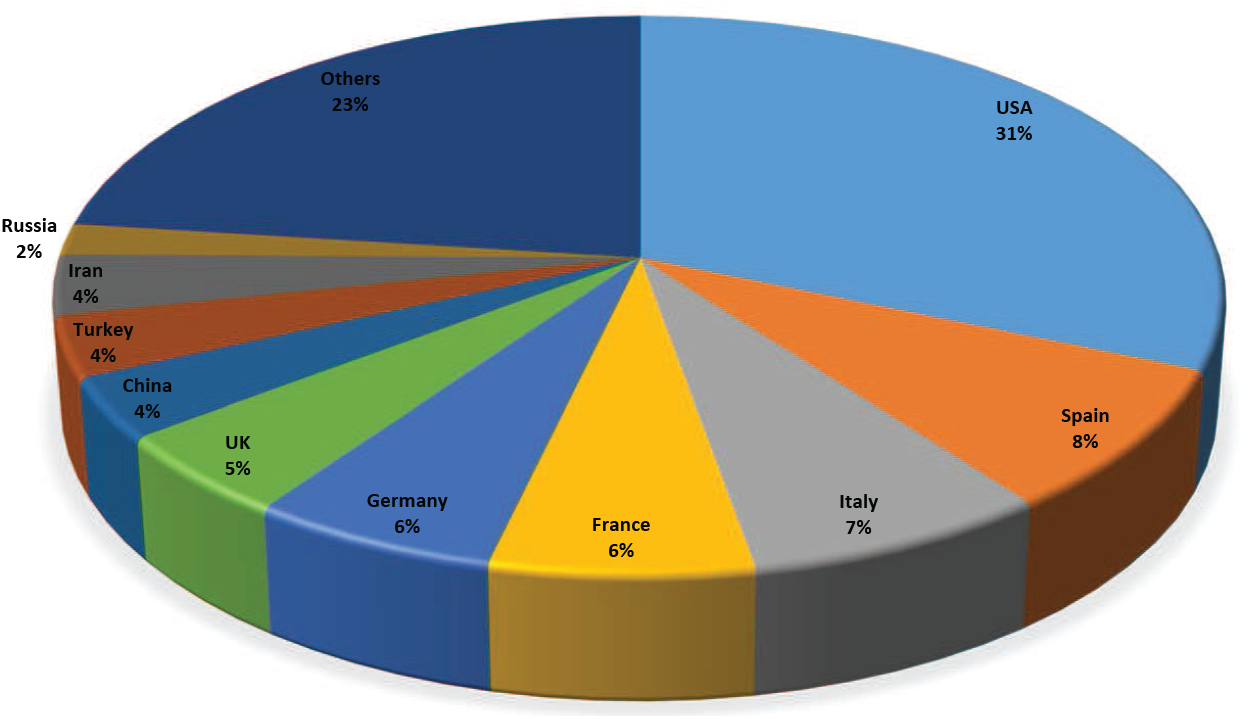
Confirmed cases of top 10 countries for accumulative COVID-19 infection till 18 April 2020.

Moreover, within the short period of time, the infected cases in USA reached to around one-third of the global confirmed cases. The global increase rate of COVID-19 infected cases over the time (till April 17, 2020) is illustrated in Fig. 3. The figure reveals that the rate increases initially due to China but the number of infected cases are not too high. However, the infected rate again increases after the second week of March, 2020 because of spreading COVID-19 globally specially in Europe, USA and Iran.

**Fig 3.**
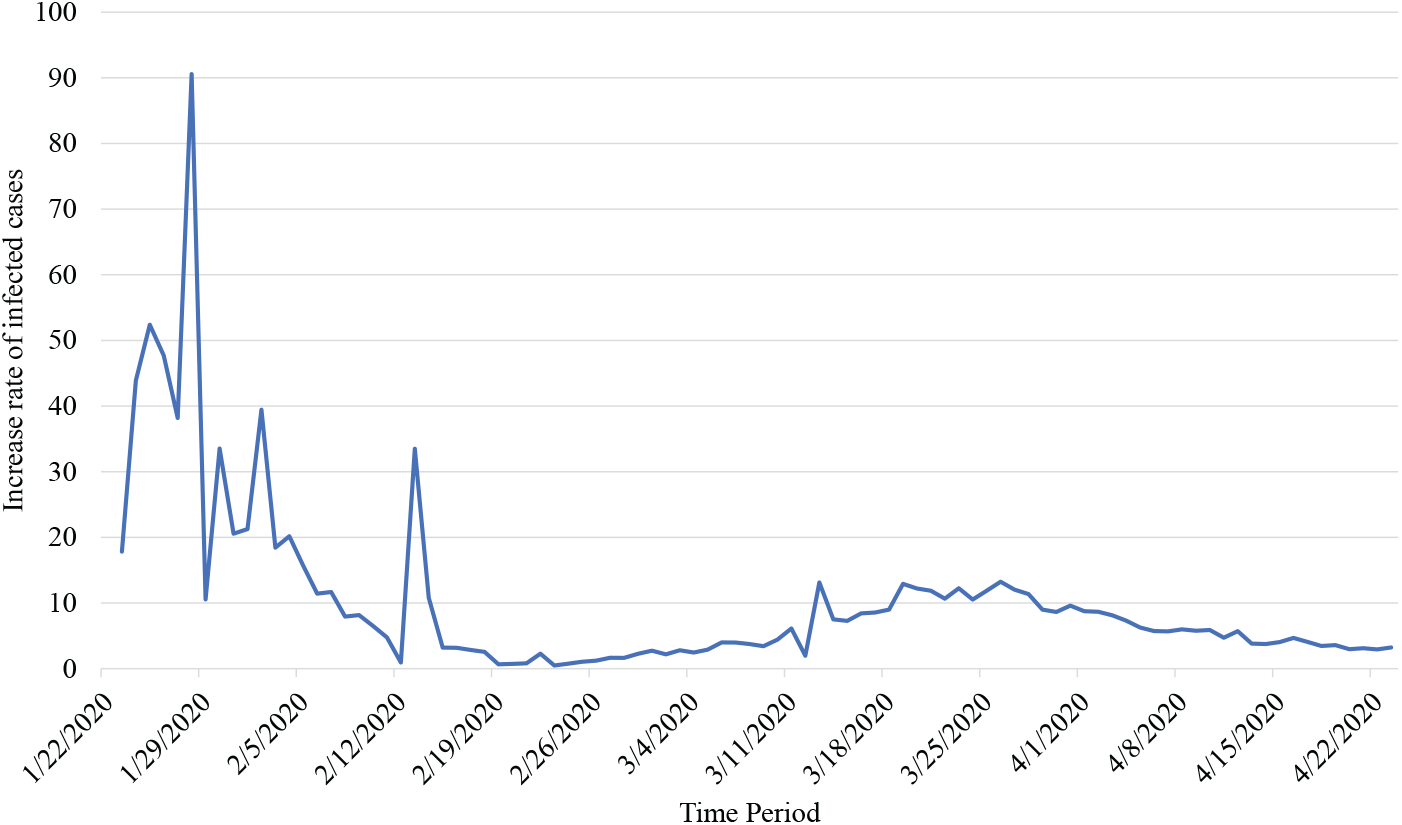
Global increase rate of COVID-19 infected cases from January 22, 2020 to April 17, 2020.

The number of human loss due to COVID-19 and also the recovered cases from COVID-19 continue to increasing throughout the time as shown in Fig. 1. The human loss due to COVID-19 is frightening in numerous countries, like USA, Italy, Spain, France, UK. The percentages of human loss for COVID-19 of top 10 countries over all the loss of the globe is shown in Fig. 4. However, only three countries namely USA, Italy and Spain contribute ore than 50% of COVID-19 deaths as presented in Fig. 4.

**Fig 4.**
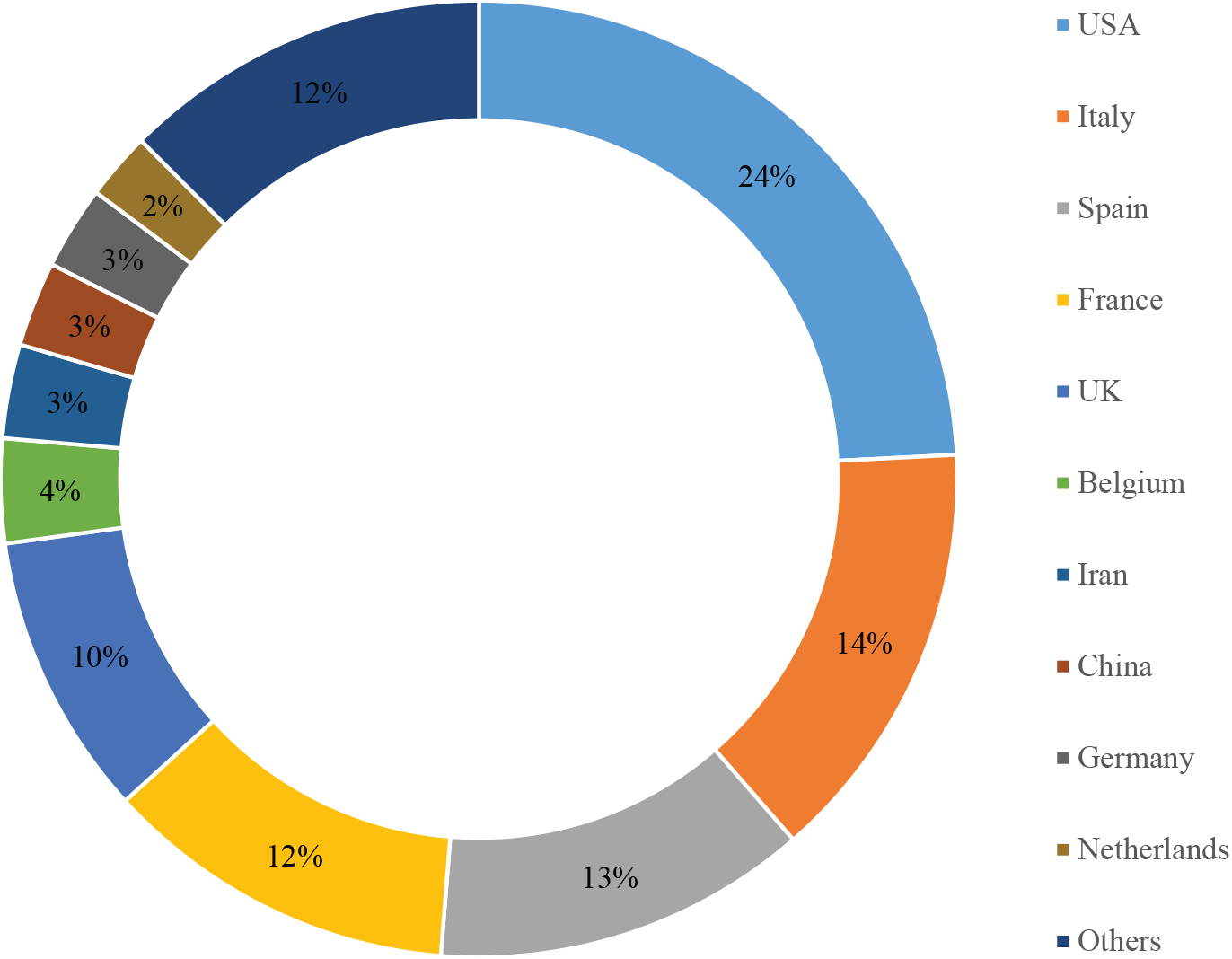
Death cases of top 10 countries due to COVID-19 till 18 April 2020.

Fig. 5 shows the gaps among infected, death, and recovery cases (COVID-19) for 10 most infected countries. This figure reveals that the gap between the infected and recovered cases is narrow for China only. Hence, China is the only country that is handling this COVID-19 crisis successfully compare to other countries. However, the differences between the death and recovered cases for most of the countries are not significant. Moreover, the death cases surpasses recovered cases for UK among these 10 countries. Fig. 6 shows the death and recovered percentages with respect to infected cases for top 10 COVID-19 infected countries to get inside the scenario. Though the average death case is 7% for these countries due to COVID-19, the ratios are significantly high for Italy, UK, France, Spain. However, despite limited knowledge regarding COVID-19, different countries are trying their best to fight against this pandemic. Fig. 6 also exhibits that the recovery cases from COVID-19 are satisfactory in China (93.14%), Iran (69.36%) and Germany (61.10%) only, other countries are still straggling to find the right ways. By the way, the average recovered rate is only 34% for these countries till now. The demographic information of the infected and death cases are presented in Fig. 7, 8, and 9. Fig. 7 shows the percentage of COVID-19 infected cases by sex based on [32] and infection among females are more. However, the death percentage for COVID-19 is more for male than female as shown in Fig. 8. Fig. 9 shows the percentages of infected and death cases among different age groups based upon [32]. The figure depicts that the infection cases are higher for ages from 21 to 70. The main cause of the characteristics is that these groups are working class and come contact with many peoples. However, the death cases are higher for ages above 60 years as shown in Fig. 9. The main cause is the weak immune system for older peoples.

**Fig 5.**
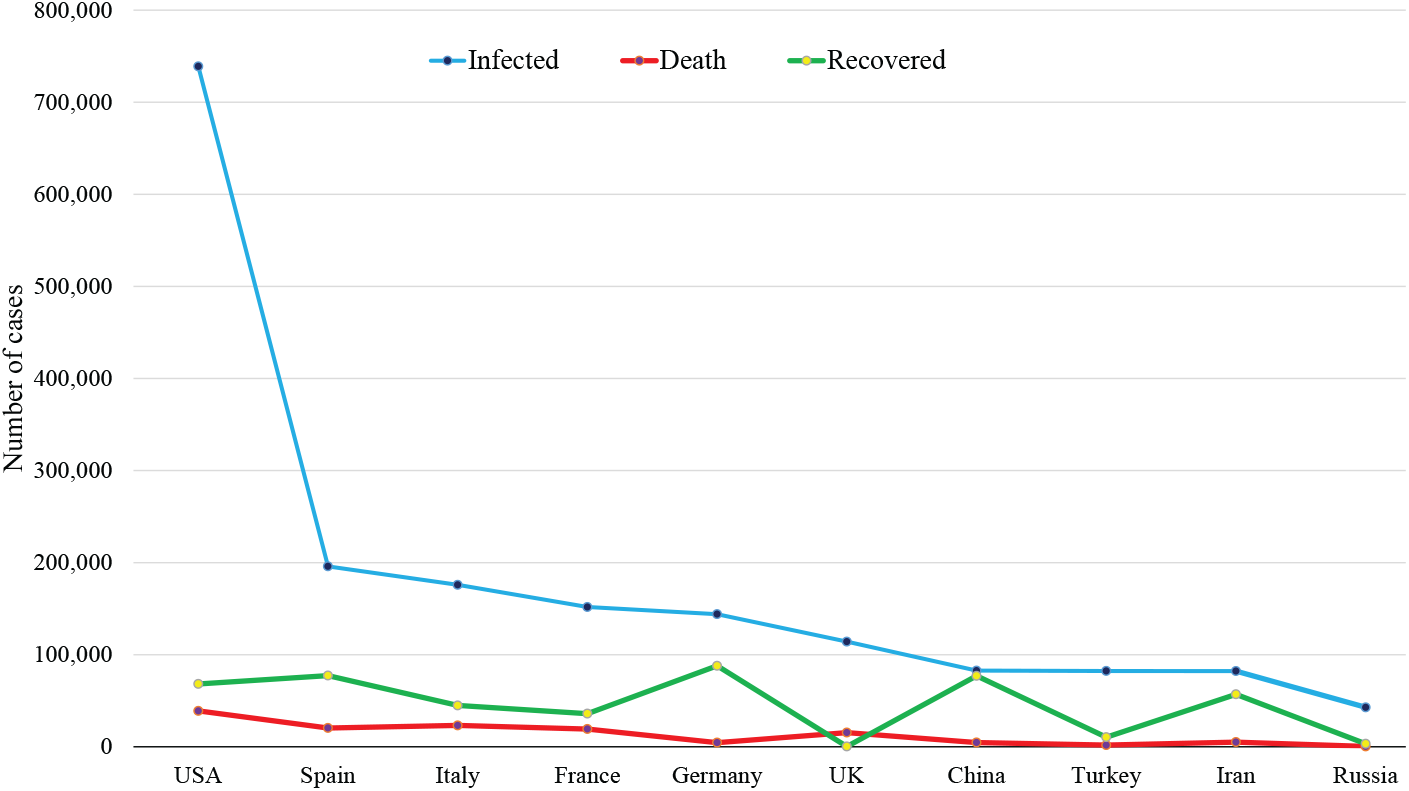
Cumulative infected, death and recovered cases of COVID-19 for top 10 infected countries till 17 April 2020.

**Fig 6.**
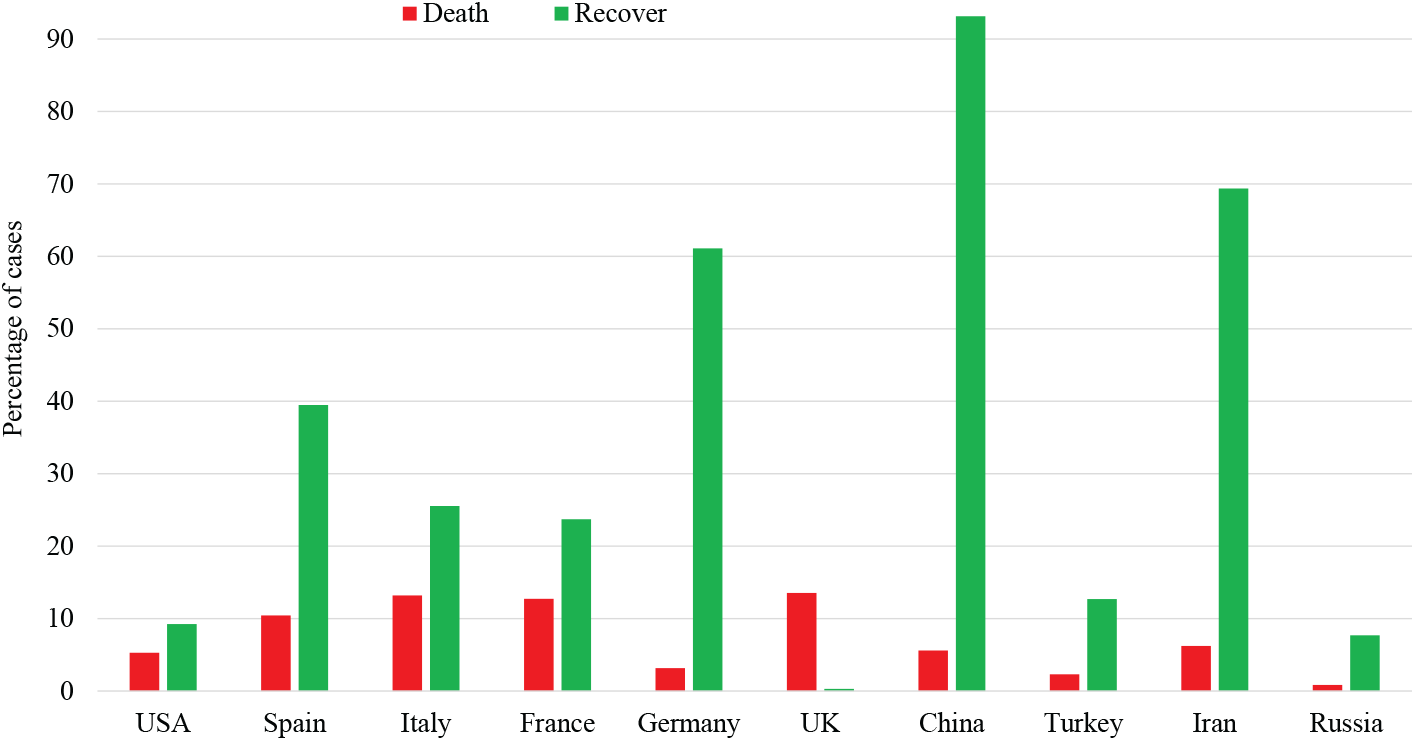
Comparison of death and recovered ratio with respect to infected cases of COVID-19 for top 20 infected countries as on April 17, 2020.

**Fig 7.**
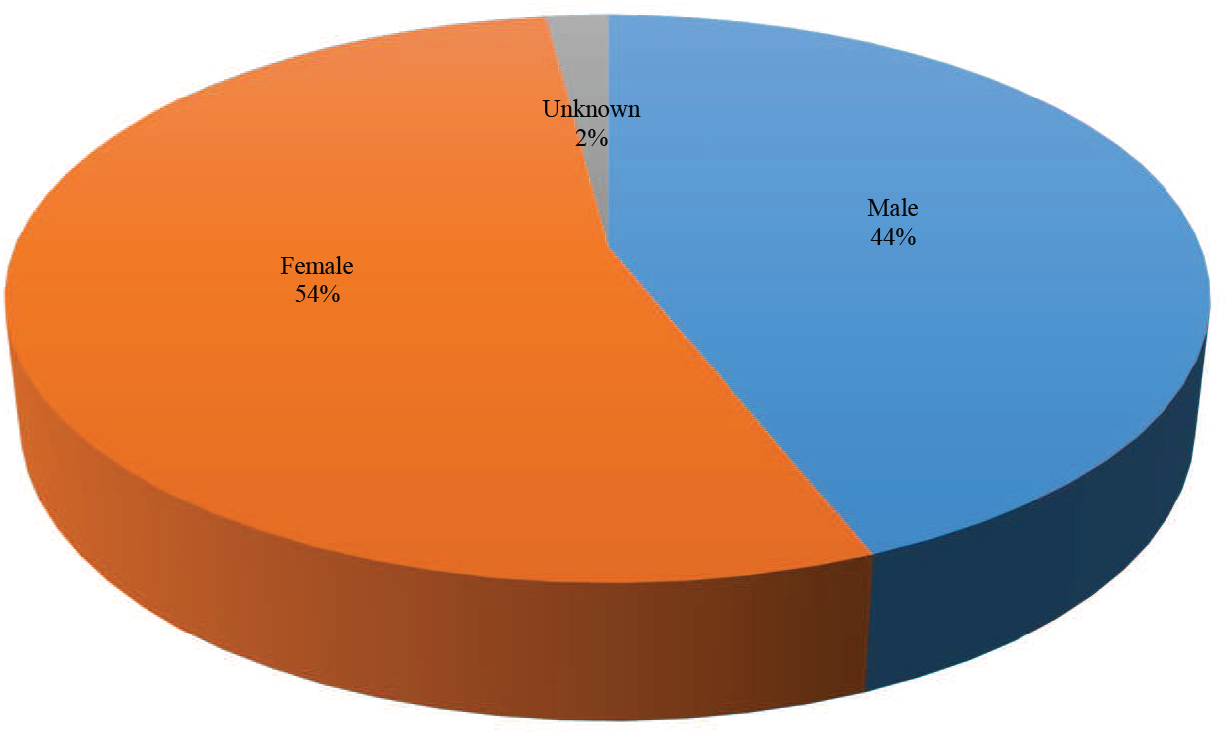
COVID-19 infected cases by sex as on April 14, 2020.

**Fig 8.**
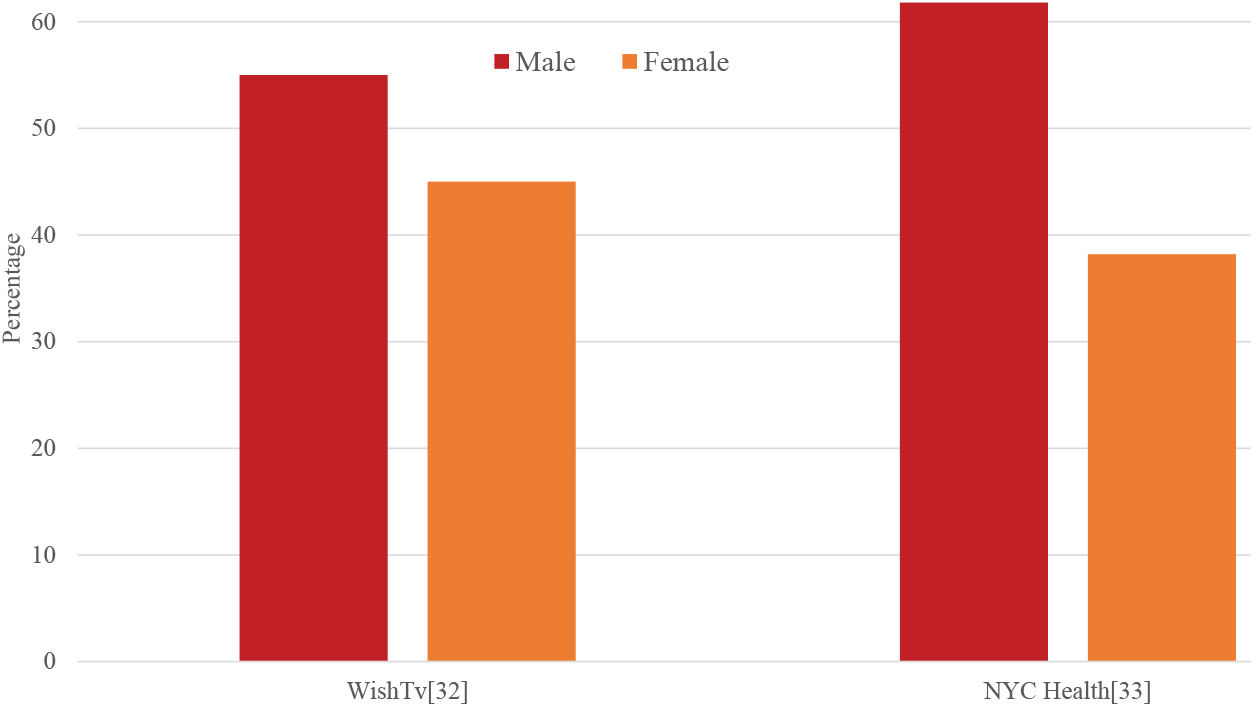
COVID-19 death cases by sex as on April 14, 2020.

**Fig 9.**
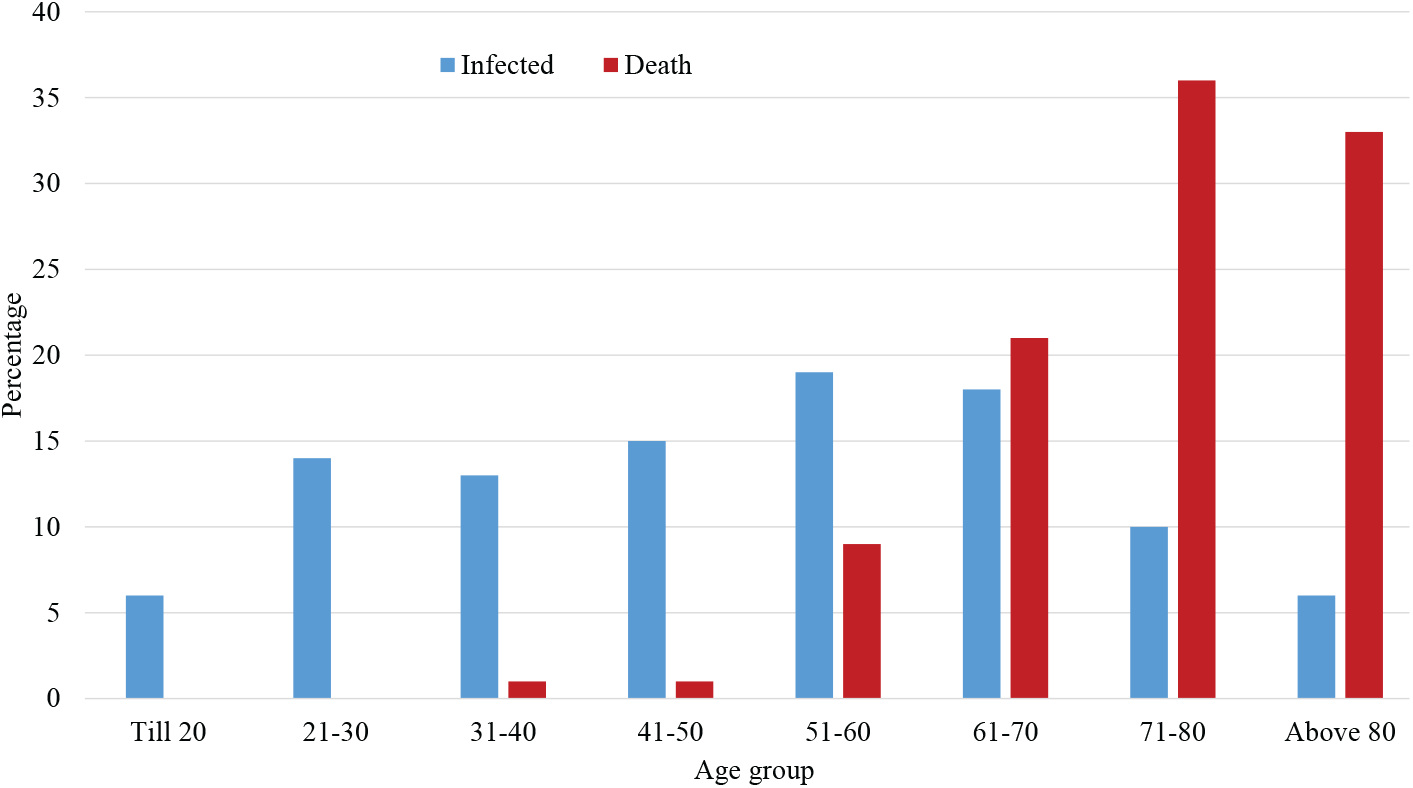
COVID-19 infected and death cases by age group as on April 14, 2020.

Fig. 10 illustrates the total COVID-19 tests performed by different countries per 1,000 people based on the data collected from Our World in Data [7]. Norway, Australia and South Korea are able to control COVID-19 epidemic by doing huge number of tests to their people among these countries.

**Fig 10.**
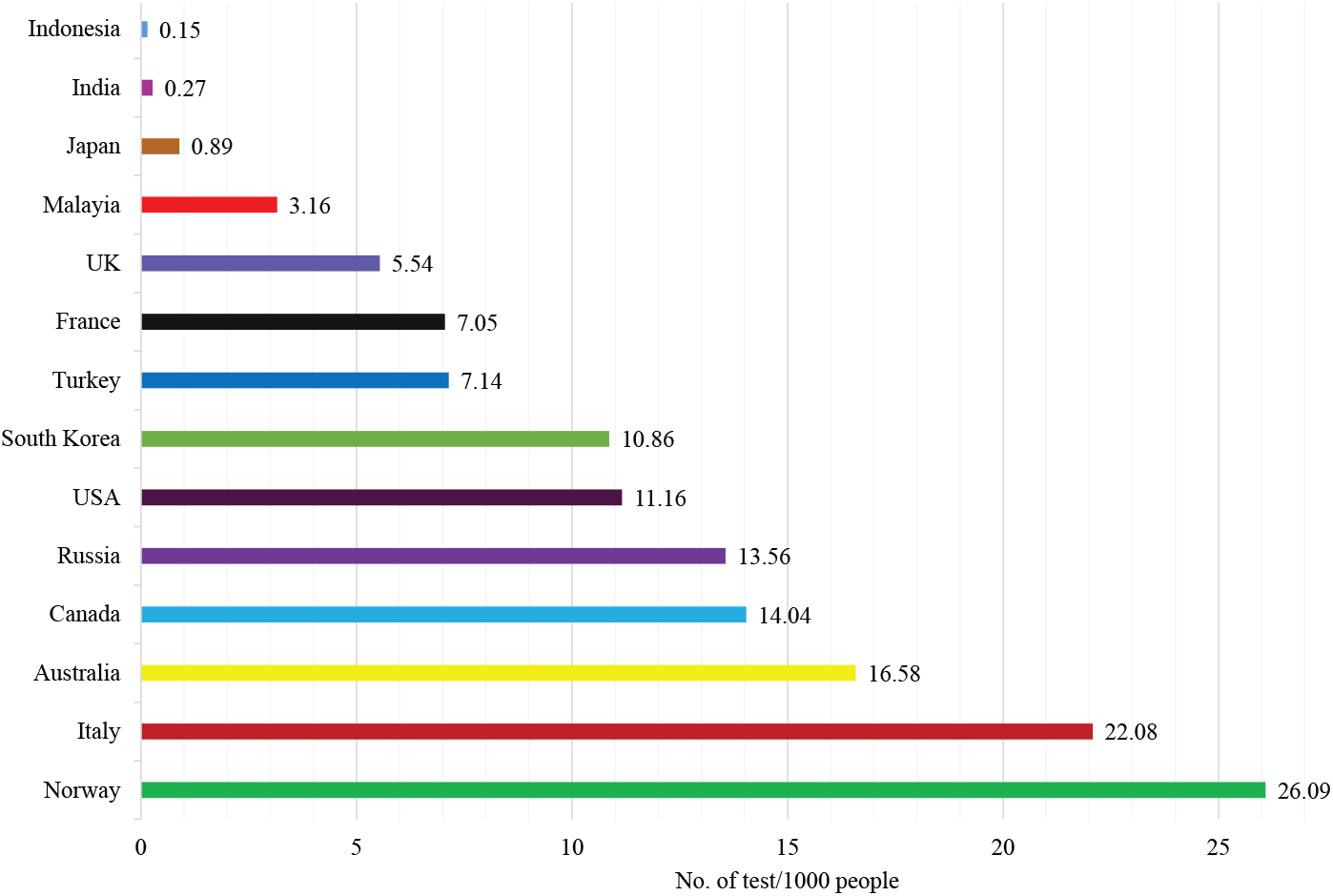
Total COVID-19 tests per 1000 people performed by countries as on April 18, 2020.

We present the predicted results for infected, recovered, death, and active cases in Fig. 11 for April 14, 2020 to May 13, 2020 based on the global data from January 22, 2020 to April 13, 2020. Here, we use vector autoregression (VAR) model to predict the cases. According to our prediction, the infected, recovered, death, and active cases of COVID-19 globally will be 3, 060, 998, 940, 380, 215, 245, and 1, 905, 372, respectively, on April 28, 2020 whereas the same values on April 13 will be 3, 521, 572, 1,485, 872, 257, 614, and 1, 778, 085, respectively. The active cases will increase as the recovered cases are increasing worldwide.

**Fig 11.**
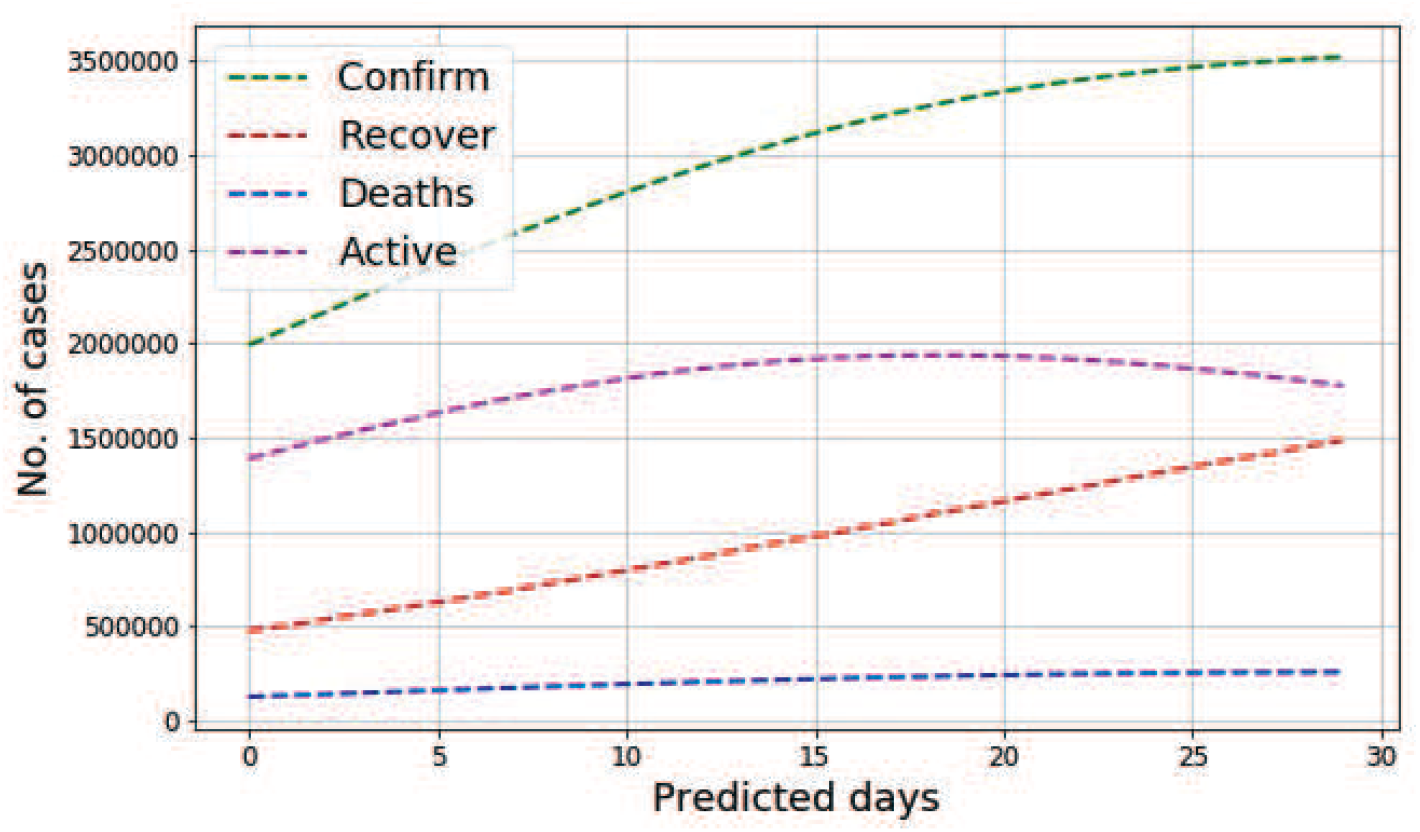
Prediction for confirm, recover, death and active cases of COVID-19 from April 14, 2020 to May 13, 2020 based on the global data from January 22, 2020 to April 13, 2020.

## 3 Literature Review

COVID-19 is the seventh coronavirus identified to contaminate humans. Individuals were first affected by the 2019-nCoV virus from bats and other animals that were sold at the seafood market in Wuhan [15, 24]. Afterward, it began to spread from human to human mainly through respiratory droplets produced while people sneeze cough or exhaling [3]. Epidemiological and/or clinical characteristics of COVID-19 are analyzed in the studies [9–14].

In [9], the authors investigate the epidemiologic and clinical characteristics based on 91 cases of COVID-19 patients of Zhejiang, China. Among these samples, 96.70% were laboratory-confirmed COVID-19 tested positive for SARS-Cov-2 while 3.30% were clinical-diagnosed COVID-19 cases. The average age of the patients was 50 while females accounted for 59.34%. The typical indications were fever (71.43%), cough (60.44%) and fatigue (43.96%). 43.96%o these patients were affected from local cases, 34.07% went to or were in Wuhan/Hubei, 8.79% came in contact with peoples from Wuhan, and 12.09% were from aircraft transmission. The authors represent a detailed statistical analysis of 1, 212 individuals collected from January 21 to February 14, 2020, and covering 18 regions of the Henan province, China [10]. Among these cases, 55% were male and ages of these patients were from 21 to 60 years. Among these patients, 20.63% had Wuhan’s travel history. In [11], the authors investigate epidemiological, demographic, clinical, and radiological features and laboratory data for 99 cases of 2019-nCoV collected from Jinyintan Hospital, Wuhan, China. They found that 49% of these patients traveled to the Huanan seafood market. The average age of the victims was 55.5 years, and most of them (67.68%) were men. The main clinical manifestations were fever (83%), cough (82%), shortness of breath (31%). Among the sufferers, 75% exhibited bilateral pneumonia also. The work in [12] analyzes the clinical characteristics of 1,099 patients with laboratory-confirmed 2019-nCoV ARD from 552 hospitals in 31 provinces/provincial municipalities of Wuhan, China. This work concluded that the median age of these patients was 47 years where 41.90% of them were female. The most common symptoms of these patients were fever (87.9%) and cough (67.7%). Most of these cases had a Wuhan connection (31.30% had been to Wuhan, and 71.80% had contacted people from Wuhan). Epidemiological investigations were conducted in [13] among all close contacts of COVID-19 patients (or suspected patients) in Nanjing, Jiangsu Province, China. Among them, 33.3% recently traveled Hubei and the average age of these cases was 32.5 years including 33.3% male. 20.8% of these patients showed fever, cough, fatigue symptoms during hospitalization whereas 50.0% cases showed typical CT images of the ground-glass chest and 20.8% presented stripe shadowing in the lungs. The study in [14] estimates the clinical features of COVID-19 in pregnancy and the intrauterine vertical transmission potential of COVID-19 infection. The age range of the subjects was 26–40 years and everybody of them had laboratory-confirmed COVID-19 pneumonia. They showed a similar pattern of clinical characteristics to non-pregnant adult patients. The authors mainly found that no intrauterine fetal infections occurred as a result of COVID-19 infection during a late stage of pregnancy.

Machine learning can play an important role to detect COVID-19 infected people based on the observatory data. The work in [16] proposes an algorithm to investigate the readings from the smartphone’s sensors to find the COVID 19 symptoms of a patient. Some commons symptoms of COVID-19 victims like fever, fatigue, headache, nausea, dry cough, lung CT imaging features, and shortness of breath can be captured by using the smartphone. This detection approach for COVID-19 is faster than the clinical diagnosis methods. The authors in [17] propose an artificial intelligence (AI) framework for obtaining the travel history of people using a phone-based survey to classify them as no-risk, minimal-risk, moderate-risk, and high-risk of being affected with COVID-19. The model needs to be trained with the COVID-19 infected information of the areas where s/he visited to accurately predict the risk level of COVID-19. In [18], the authors develop a deep learning-based method (COVNet) to identify COVID-19 from the volumetric chest CT image. For measuring the accuracy of their system, they utilize community-acquired pneumonia (CAP) and other non-pneumonia CT images. The authors in [19] also use deep learning techniques for distinguishing COVID-19 pneumonia from Influenza-A viral pneumonia and healthy cases based on the pulmonary CT images. They use a location-attention classification model to categorize the images into the above three groups. Depth cameras and deep learning are applied to recognize unusual respiratory pattern of personnel remotely and accurately in [20]. They propose a novel and effective respiratory simulation model based on the characteristics of original respiratory signals. This model intends to fill the gap between large training datasets and infrequent real-world data. Multiple retrospective experiments were demonstrated to examine the performance of the system in the detection of speculated COVID-19 thoracic CT characteristics in [21]. A 3D volume review, namely “Corona score” is employed to assess the evolution of the disease in each victim over time. In [22], the authors use a pre-trained UNet to fragment the lung region for automatic detection of COVID-19 from a chest CT image. Afterward, they use a 3D deep neural network to estimate the probability of COVID-19 infections over the segmented 3D lung region. Their algorithm uses 499 CT volumes as a training dataset and 131 CT volumes as a test dataset and achieves 0.959 ROC AUC and 0.976 PR AUC. The study in [23] presents evidence of the diversity of human coronavirus, the rapid evolution of COVID-19, and their clinical and Epidemiological characteristics. The authors also develop a deep learning model for identifying COVID-19. and trained the model using a small CT image datasets. They find an accuracy of around 90% using a small CT image dataset.

In [25], the authors propose a stochastic transmission model for capturing the phenomenon of the COVID-19 outbreak by applying a new model to quantify the effectiveness of association tracing and isolation of cases at controlling a severe acute respiratory syndrome coronavirus 2 (SARS-CoV-2)-like pathogen. In their model, they analyze synopses with a varying number of initial cases, the basic reproduction number, the delay from symptom onset to isolation, the probability that contacts were traced, the proportion of transmission that occurred before symptom start, and the proportion of subclinical infections. They find that contact tracing and case isolation are capable enough to restrain a new outbreak of COVID-19 within 3 months. In [37], the author points out some of the features of COVID-19 pandemic with the help of Prisoner’s dilemma. However, the work in [37] does not provide any concrete utility functions for the studied game.

The works [9–14,16–22] focused on COVID-19 detection and analyzed the characteristic of its respiratory pattern. Hence, the literature has achieved a significant result in terms of post responses. In fact, it is also imperative to control the epidemic of COVID-19 by maintaining social distance. Therefore, different from the existing literature, we focus on the design of a model that can measure individual’s isolation and social distance to prevent the epidemic of COVID-19. The model considers both isolation and social distancing features of individuals to control the outbreak of COVID-19.

## 4 System Model and Problem Formulation

Consider an area in which a set 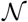 of *N* individuals are living under COVID-19 threat and must decide whether to stay at home or go leave their homes to visit a market, shop, train station, or other locations, as shown in Fig. 12. Everyone has a mobile phone with GPS. From analyzing the GPS information, we can know their home locations of each individuals, and longitude and latitude of these locations are denoted by ***X***^h^, and ***Y***^h^, respectively. We consider one time period (e.g., 15 or 30 minutes) for our scenario and this time period is divided into *T* smaller time steps in a set 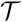. For each of time step 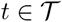, we have the GPS coordinates ***X*** and ***Y*** of every individual.

**Fig 12.**
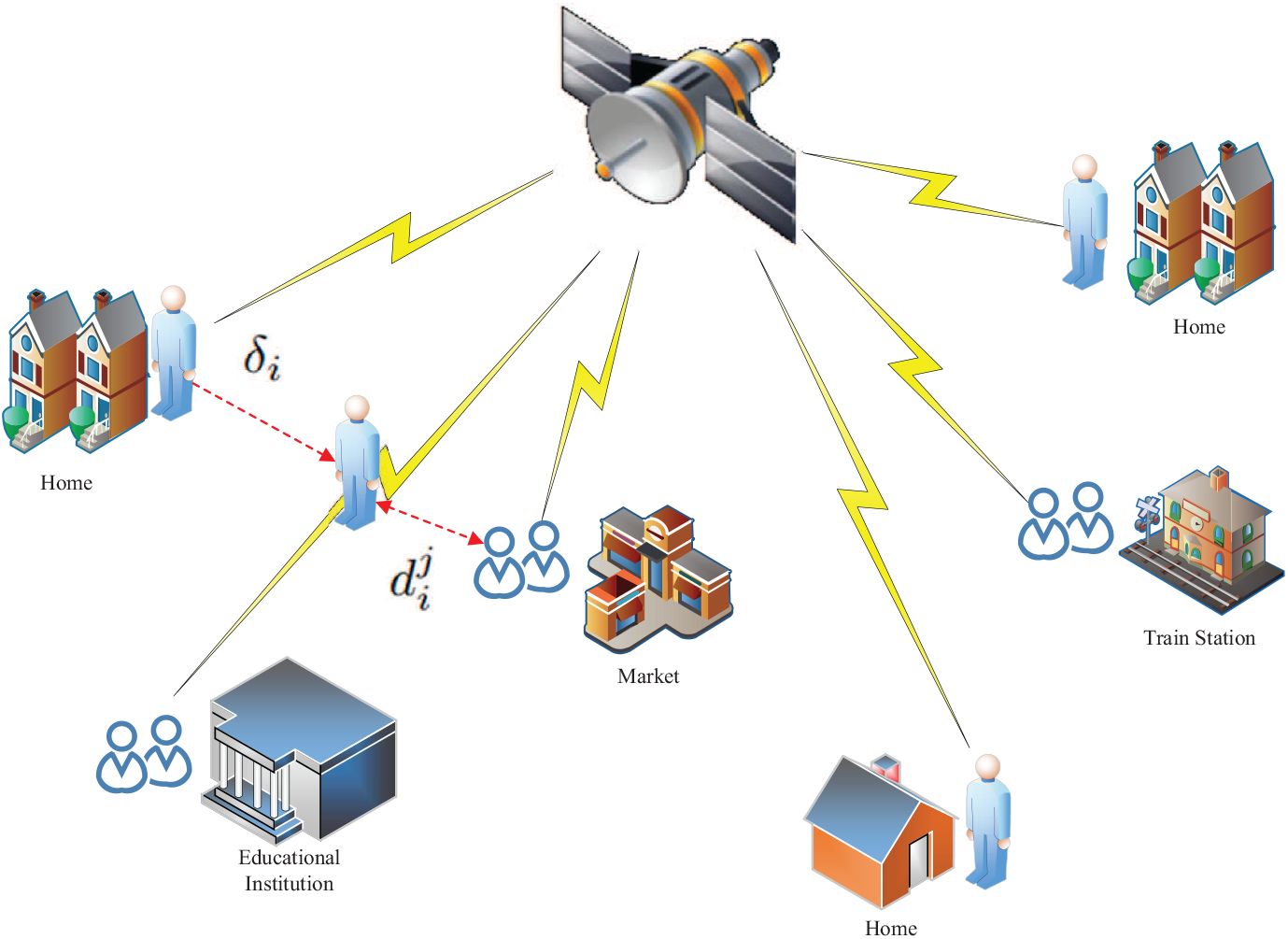
Exemplary System model. Isolation indicates staying at home whereas social distancing measures the distance of a individual from others.

Now, the deviation from home for any individual 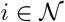 in between two time steps can be measured by using Euclidean distance as follows:

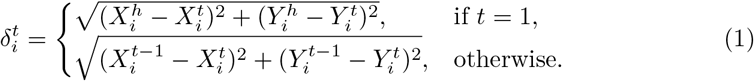

Thus, the total deviation from home by each individual 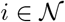 in a particular time period can be calculated as follows:

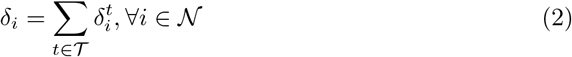

Hence, the grand deviation of all the individuals of 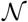 can be summarized as follows:

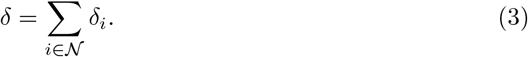

On the other hand, at the end of a particular time period, the distance between an individual 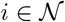 and any other individuals 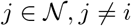 is as follows:

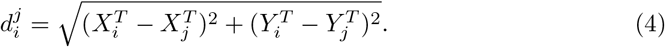

Hence, the total distance of individual 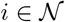 from other individuals 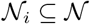, who are in close proximity with 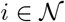, can be expressed as follows:

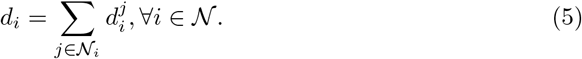

In the similar fashion of (3), the grand distance of all 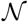 individual can be summarized as follows:

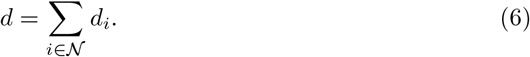

Our objective is to keep *δ* minimum for reducing the spread of COVID-19 from infected individuals, which is an isolation strategy. Meanwhile, we want to maximize social distancing which mathematically translates into maximizing *d* for reducing the chance of infection from others. However, we can use log term to bring fairness [26,29] in the objective function among all individuals. Hence, we can pose the following optimization problem:

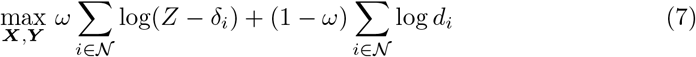

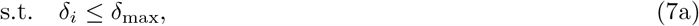

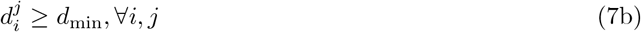

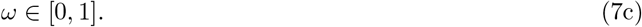

In (7), *Z* is a large number and *Z* > *δ_i_*, 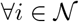. The optimization variables ***X*** and ***Y*** indicate longitude, and latitude, respectively, of the individuals. Moreover, the first term in (7) encourages individual for *isolation* whereas the second term in (7) encourages individual to maintain fair *social distancing*. In this way, solving (7) can play a vital role in our understanding on how to control the spread of COVID-19 among vast population in the society. Constraint (7a) guarantees small deviation to maintain emergency needs, while Constraint (7b) assures a minimum fair distance among all the individuals to reduce the spreading of COVID-19 from one individual to another. Constraint (7c) shows that *ω* can take any value between 0 and 1 which captures the importance between two key factors captured in the objective function of (7). For example, if COVID-19 is already spreading in a given society, then most of the weight would go to isolation term rather than social distancing. The objective of (7) is difficult to achieve as it requires the involvement and coordination among all the *N* individual. Moreover, if the individuals are not convinced then it is also difficult for the government to attain the objective forcefully. Thus, we need an alternative solution approach that encourage individual separately to achieve the objective and game theory, which is successfully used in [27,28], can be one potential solution, which will be elaborated in the next section.

## 5 A Noncooperative Game Analysis for Achieving Social Objective

To attain the objective for a vast population, governments can introduce incentives for isolation and also for social distancing. Then every individual wants to maximize their utilities or payoffs. In this way, government can play its role for achieving social objective. Hence, the modified objective function is given as follows:

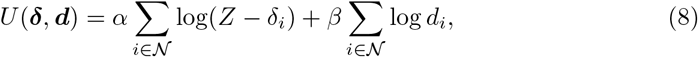

where *α* = *α′ω* and *β* = *β′* (1 − *ω*) with *α′* > 0 and *β′* > 0 are incentives per unit of isolation and social distancing. In (8), one individual’s position affects the social distancing of others, and hence, the individuals have partially conflicting interest on the outcome of *U*. Therefore, the situation can be explained with the noncooperative game [35,36].

A noncooperative game is a game exhibiting a competitive situation where each player needs to make it’s choice independent of the other players, given the possible policies of the other players and their impact on the player’s payoffs or utilities. Now, a noncooperative game in strategic form or a strategic game 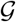 is a triplet 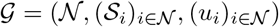 [30] for any time period where:

- 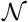 is a finite set of players, i.e., 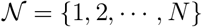,
- 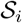 is the set of available strategies for player 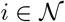,
- 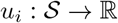 is the payoff function of player 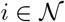, with 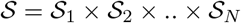.

In our case 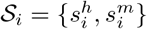 where 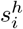 and 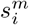 indicate the strategies of staying at home and moving outside for player 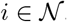, respectively. The payoff or incentive function of any player 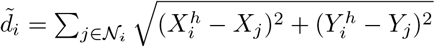 in a time period can be defined as follows:

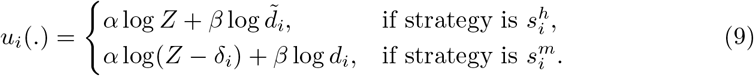

where 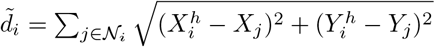.

The Nash equilibrium [31] is the most used solution concept for a noncooperative game. Formally, Nash equilibrium can be defined as follows [34]:

### Definition 1

*: A pure strategy Nash equilibrium for a non-cooperative game 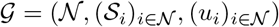 is a strategy profile* 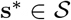 *where 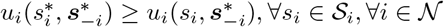*.

However, to find the Nash equilibrium, the following two definitions can be helpful.

### Definition 2

*[30]: A strategy* 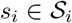 *is the dominant strategy for player* 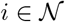 *if* 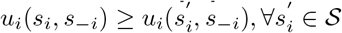 *and* 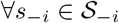, *where* 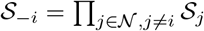 *is the set of all strategy profiles for all players except i*.

### Definition 3

*[30]: A strategy profile* 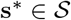 *is the dominant strategy equilibrium if every elements* 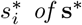 *is the dominant strategy of player* 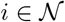.

Thus, if we can show that every player of our game 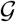 has a strategy that gives better utility irrespective of other players strategies, then with the help of Definition 2 and 3. Our game exhibits the structure of an N-player Prisoner’s dilemma, so based on [30] it will have a unique Nash equilibrium.

Thus, Nash equilibrium is the solution of the noncooperative game 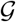. In this equilibrium, no player of 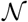 has the benefit of changing their strategy while others remain in their strategies. That means, the utility of each player 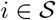 is maximized in this strategy, and hence ultimately maximize the utility of (8). To this end, maximizing *U* of (8) ultimately maximize the original objective function of (7).

## 6 Sustainability of Lockdown Policy with the System Model

The total amount of incentive a particular time period is presented in (8). In a particular day, we have 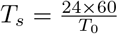 time period where *T*_0_ is the length of a time period in minutes. Thus, we can denote the incentive of a time stamp *t_s_* in a particular day *p* as follows:

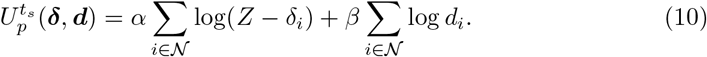

Hence, the incentive in a particular day, *p* can be given as follows:

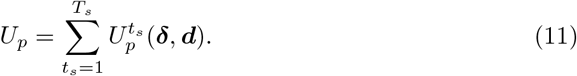

Now, if we are interested to find the sustainability of lockdown policy for a particular country till a particular number of days, denoted by *P*, we have to satisfy the following inequality:

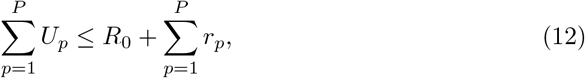

where *R*_0_ is the initial resource of a particular country starting of the lockdown policy and *r_p_* is the collected resources in a particular day, *p*, of the lockdown period. Here, *r_p_* includes governmental revenue, donation from different individuals, organizations and even countries. Moreover, the unit of *α*, *β*, *R*_0_ and *r_p_* are same.

If we assume for simplicity that *U_p_* and *r_p_* are same for every day and they are denoted by *Ũ* and 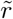, respectively, then we can rewrite (12) as follows:

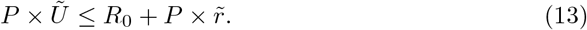

Hence, if we are interested to find the upper limit of sustainable days for a particular country using lockdown policy, then we have the following equality:

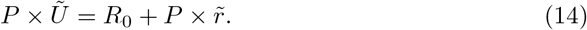

Thus, by simplifying (14), we have the following:

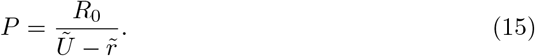

Here, the sustainable days *P* depends on *R*_0_, *Ũ*, and 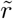. However, we cannot change *R*_0_ but government can predict 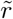. Moreover, depending on *R*_0_ and 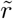, government can formulate its policy to set *α* and *β* so that individuals are encouraged to follow the lockdown policy. Alongside, we cannot continue lockdown policy infinitely based upon the limited total resources. Hence, the governments should formulate and update its lockdown policy based on the predicted sustainable capability to handle COVID-19, otherwise resource crisis will be a further bigger pandemic in the world.

## 7 Numerical Analysis

In this section, we assess the impact of the lockdown policy to control the COVID-19 outbreak using numerical analyses. We consider an area of 1, 000 m × 1, 000 m for our analysis where individuals’ position are randomly distributed. The value of the principal simulation parameters are shown in the Table 1.

**Table 1.** Value of the main simulation parameters

| Symbol | Value |
| --- | --- |
| $N$ | $\{500, 1000, 1500, 2000\}$ |
| $\alpha$ | 3.0 |
| $\beta$ | 1.0 |
| $Z$ | 1400 |
| $R_0$ | $\{5E + 23, 5.5E + 23, 6E + 23, 6.5E + 23, 7E + 23\}$ |

Fig. 13 illustrates a comparison between home isolation (stay at home) and random location in the considered area for a varying value of *ω*. In this figure, we consider two cases of *N* = 500 and *N* = 1, 000. In both the cases, home isolation (quarantine) is beneficial over staying in random location and the differences between two approaches are increased with the increasing value of *ω*. Moreover, the difference of payoffs between two approaches are increased with the increasing value of *ω* as the more importance are given in home isolation.

**Fig 13.**
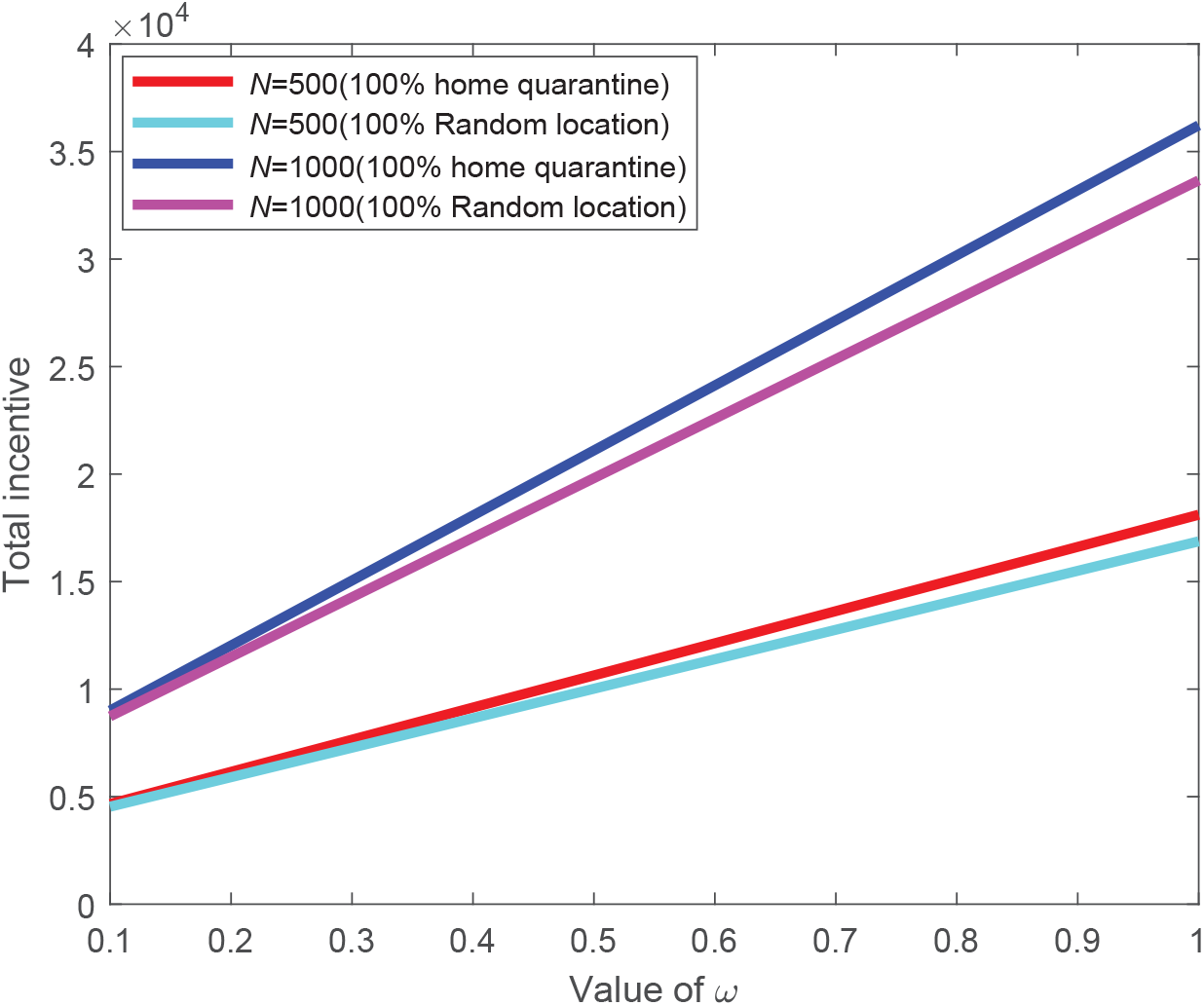
Comparison of incentive (in log scale) for varying value of *ω*.

Fig. 14,15,16,17 show the empirical cumulative distribution function (ecdf) of incentives for 500, 1,000, 1, 500, and 2,000 individuals, respectively. These figures revel that the incentive values increase with the increasing number of home quarantine individuals in all the four cases. Fig. 14 exhibits that the incentives are below 19,000, and 20, 000 for 50%, and 48% sure, respectively, for 25% and 50% home quarantine cases whereas the incentives are 90% sure in between 20, 500 and 21, 000 for 75% home isolation case. Moreover, the same values are at least 21, 500 for 50% sure in case of full home isolation. Fig. 15 depicts that the incentive of being below 38, 000 is 40% sure for 25% home isolation case, however, the same values of being above 40, 000, and 41, 000 are 40%, and 60%, sure, respective, for 50%, and 75% cases. Moreover, for 100% home isolation case, the values are in between 42, 000 to 43, 000 for sure. The incentives for 25%, 50%, 75%, and 100% home isolation cases are above 57, 000, 59,000, 61,000, and 63,000, respectively, with probability 0.60, 0.65, 0.65, and 0.80, respectively, as shown in Fig. 16. Additionally, the same values are at least 77,000, 79, 000, 81,000, and 83, 500 with 0.50, 0.50, 0.72, and 1.00 probabilities, respectively, which is presented in Fig. 17.

**Fig 14.**
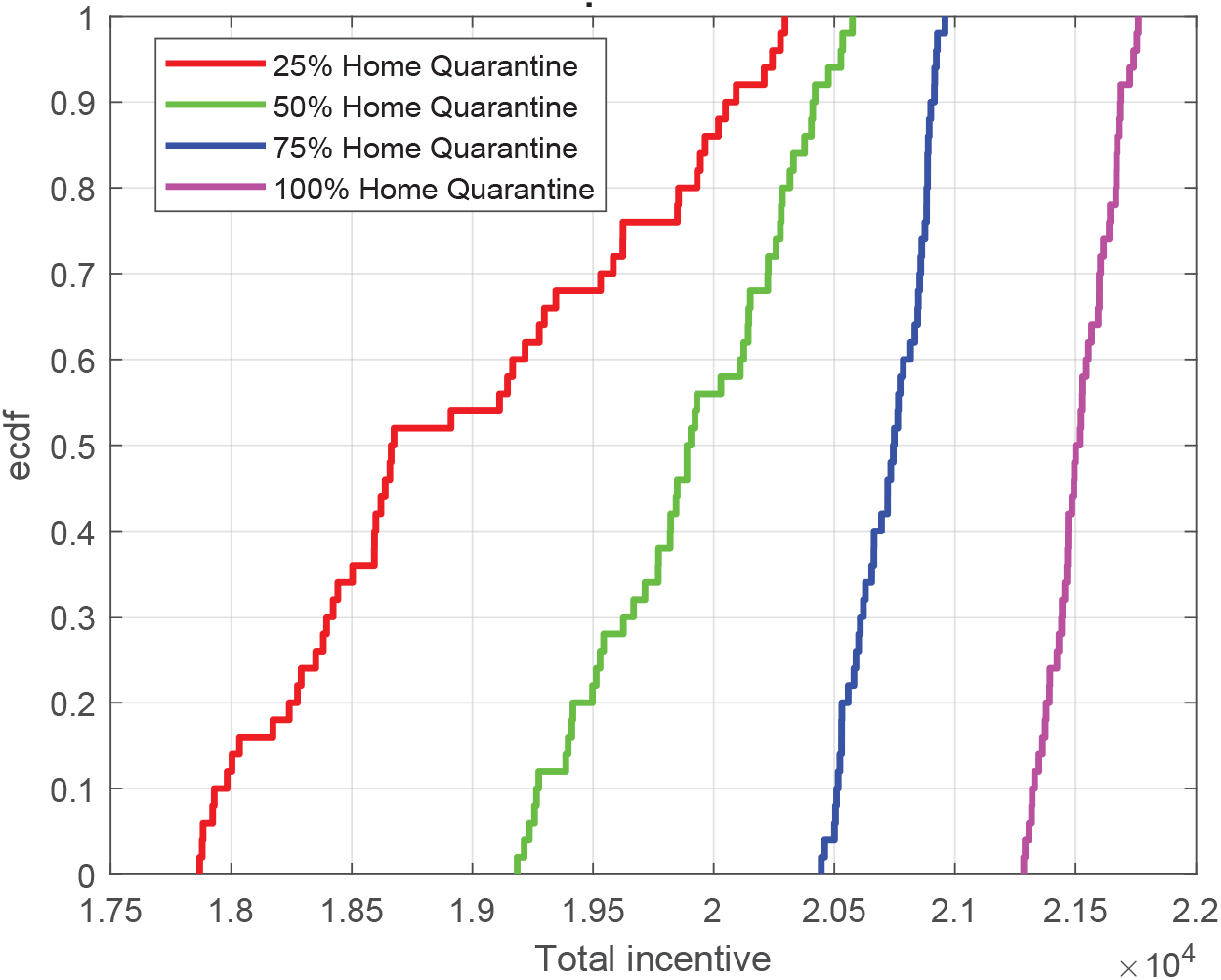
Ecdf of incentives (in log scale) for *N* = 500 with *α* = 3.0 and *β* =1.0 using 50 runs.

**Fig 15.**
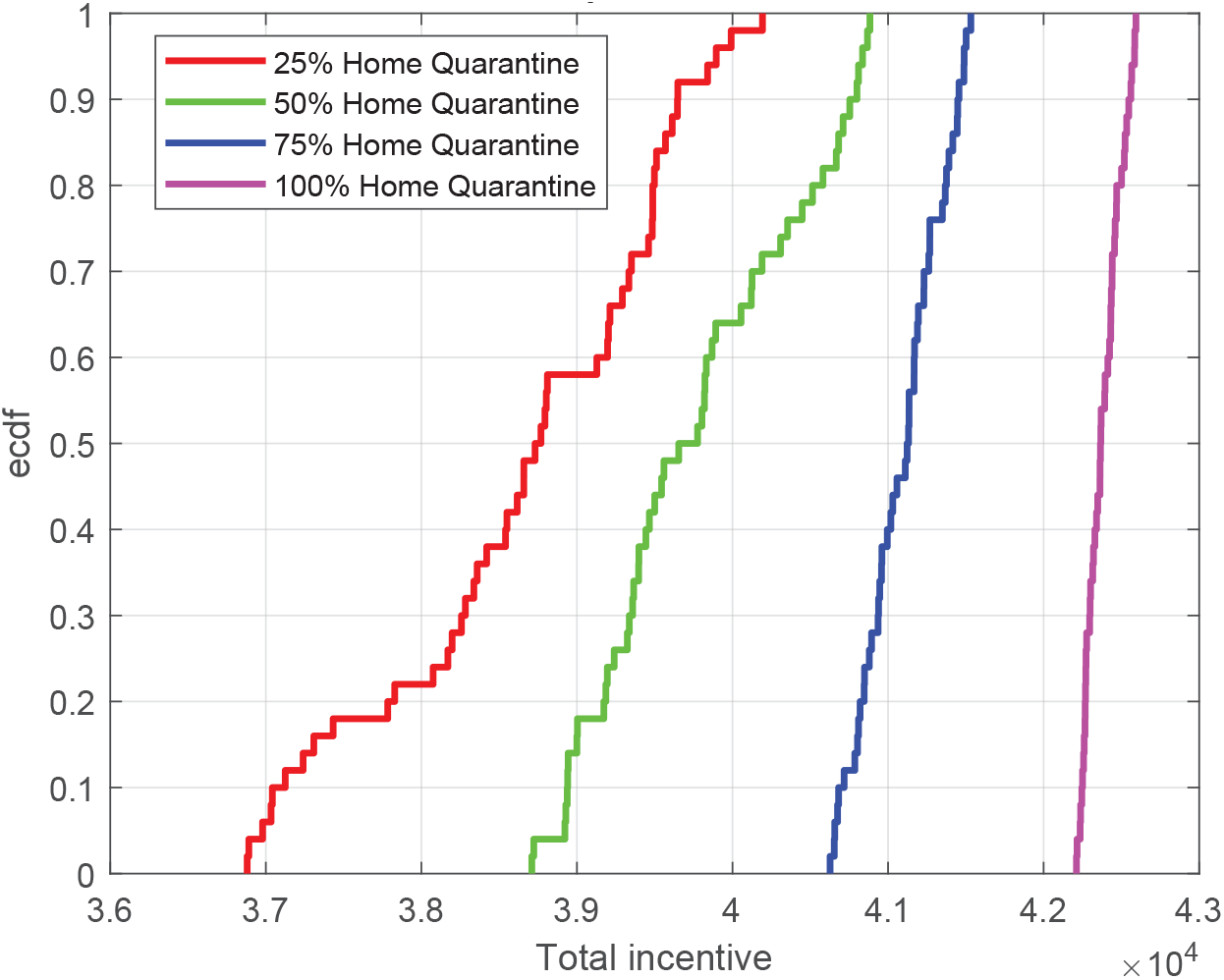
Ecdf of incentives (in log scale) for *N* = 1,000 with *α* = 3.0 and *β* = 1.0 using 50 runs.

**Fig 16.**
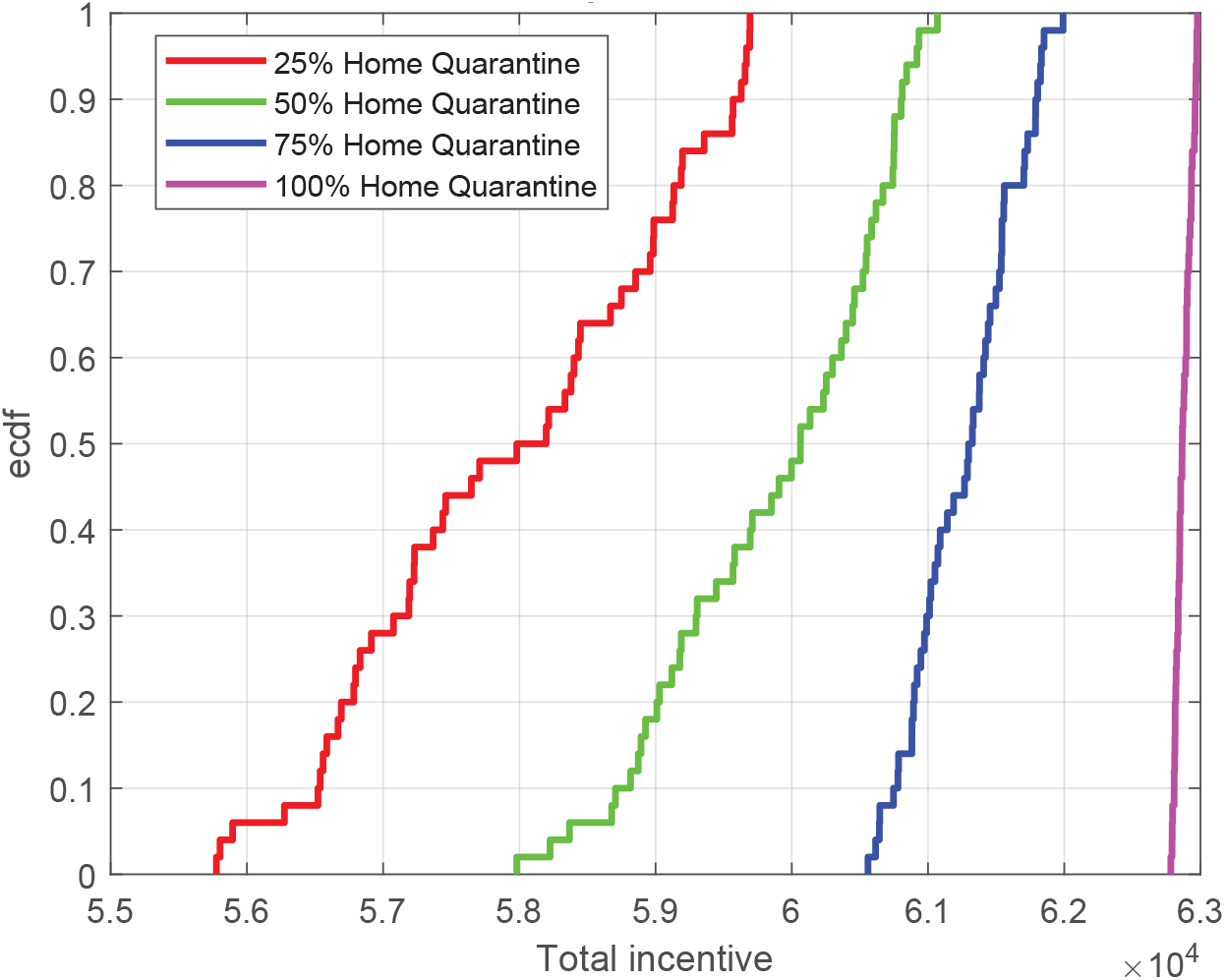
Ecdf of incentives (in log scale) for *N* = 1,500 with *α* = 3.0 and *β* = 1.0 using 50 runs.

**Fig 17.**
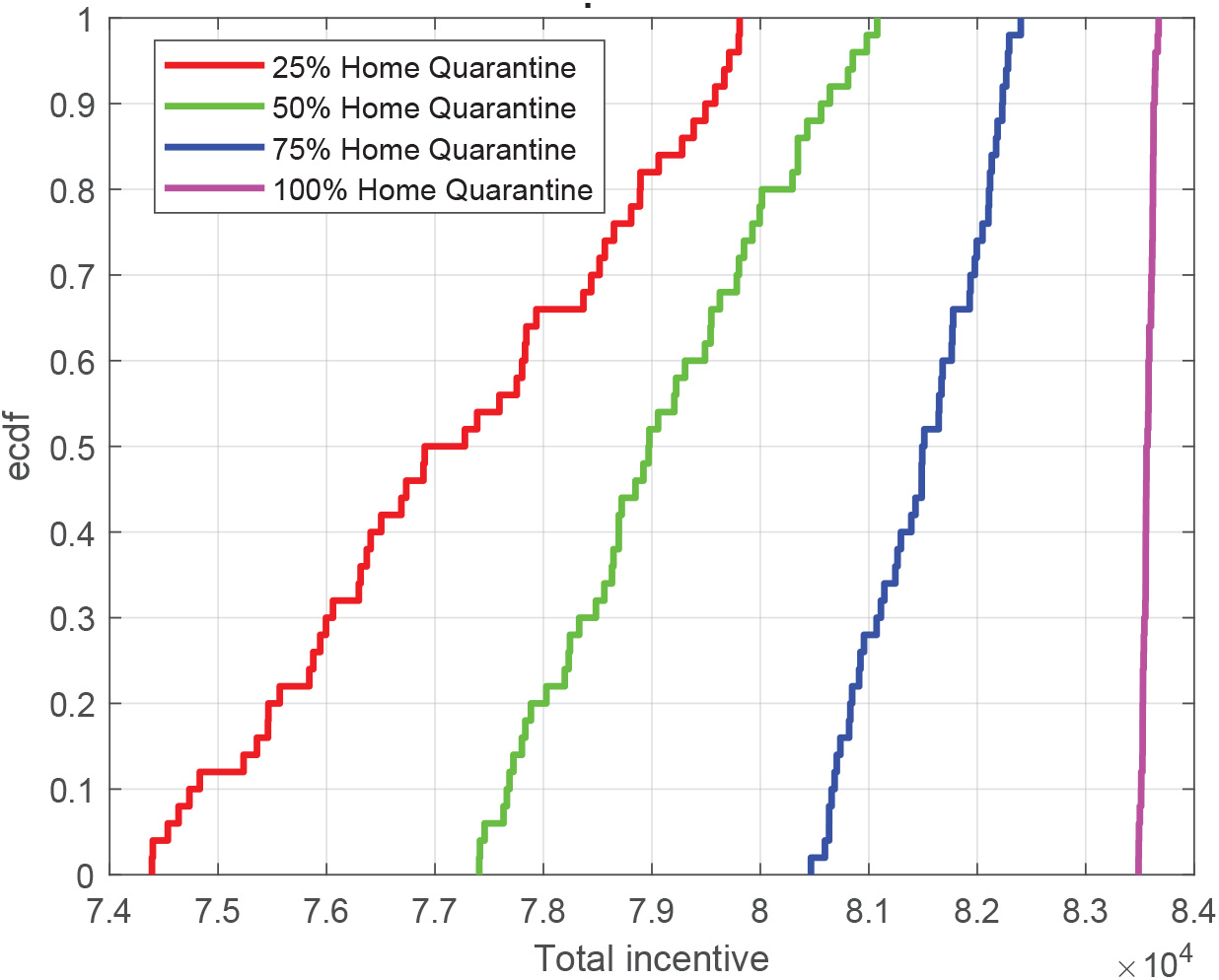
Ecdf of incentives (in log scale) for *N* = 2,000 with *α* = 3.0 and *β* =1.0 using 50 runs.

The total incentive (averaging of 50 runs) for varying percentage of home isolation individuals with different sample size are shown in Fig. 18. From this figure, we observe that the total payoff increases with increasing number of home isolation individuals for all considered cases. The incentives are 578%, 571%, 571%, and 571% better from home quarantine of 25% to 100% for *N* = 500, *N* = 1,000, *N* = 1, 500, and *N* = 2,000, respectively. Moreover, for a particular percentage of home isolation, the total incentive is related with the sample size. In case of 50% individuals in the home isolation, the incentive for *N* = 2,000 is 97.08%, 42.50%, and 15.96% more than that of *N* = 500, *N* = 1, 000, and *N* = 1, 500, respectively. Fig. 19 shows the average individual payoff for varying parentage of home isolation individuals for different scenarios. The figure exhibits that the average individual incentive increases with an increasing percentage of home isolation as the deviation *δ* decreases and hence, the value of home isolation incentive increases. For *N* = 500, the incentive of 100% home isolation is 85.25% more than that of 25% home isolation. Moreover, in a particular percentage of home isolation, the incentive decreases with an increasing number of considered individuals as the social distancing decreases due to the more number of individuals. In case of 50% home isolation, the individual incentive for *N* = 500 is 102.96% more than that of *N* = 2,000.

**Fig 18.**
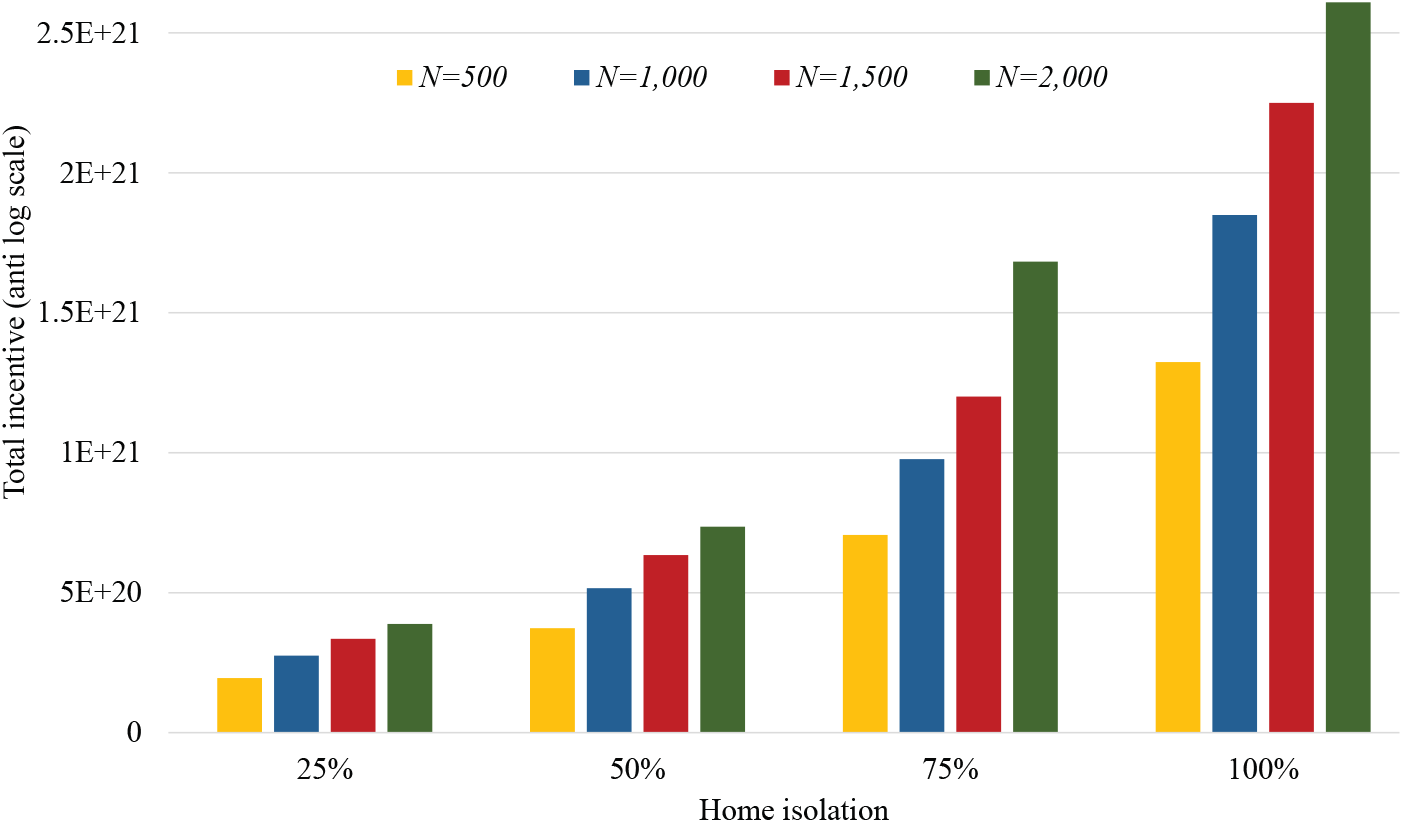
Total incentive (average of 50 runs) for varying percentage of home isolation individuals when *α* = 3.0 and *β* = 1.0.

**Fig 19.**
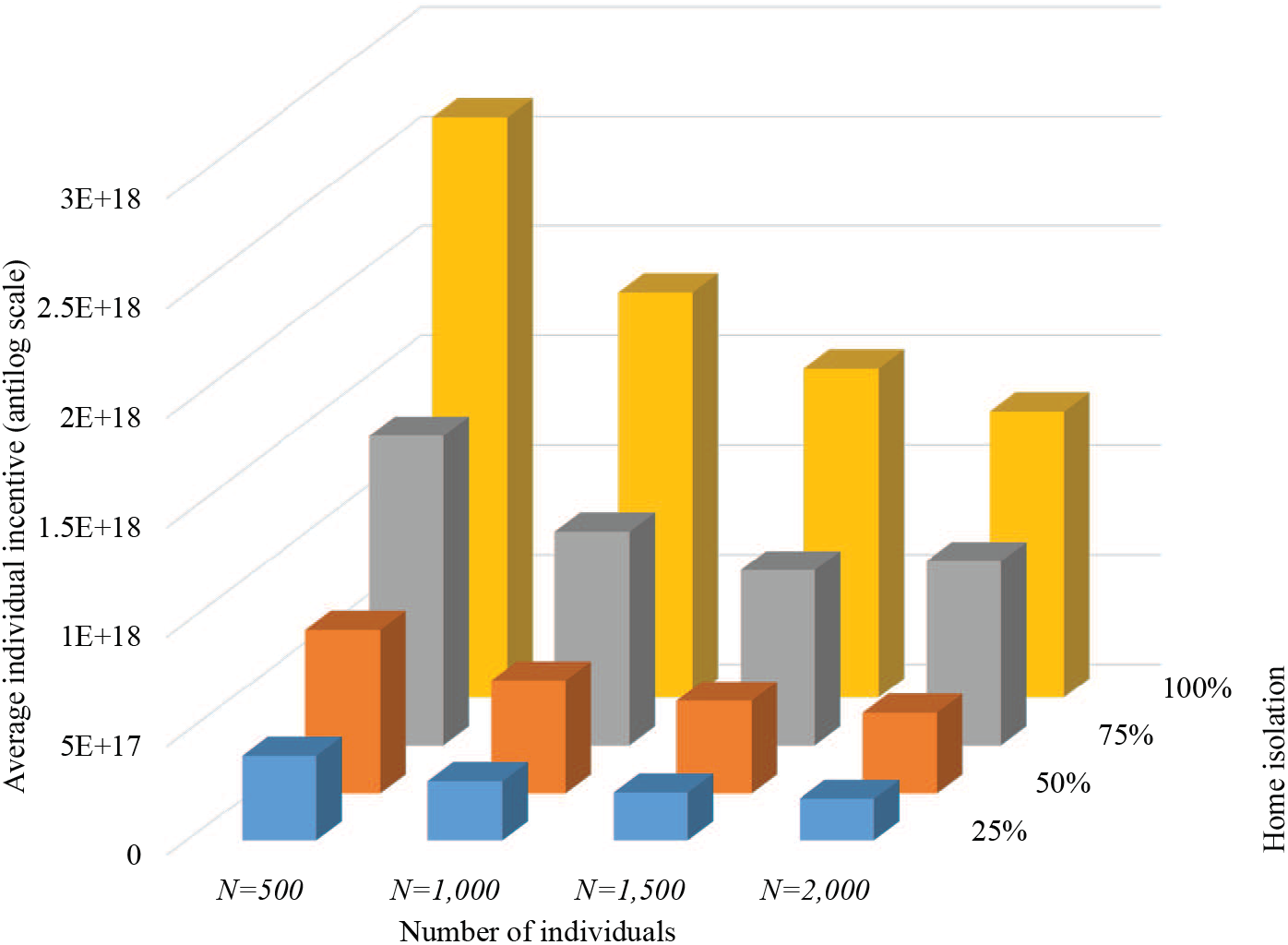
Average individual incentive for varying percentage of home quarantine individuals when *α* = 3.0 and *β* = 1.0.

Fig. 20 shows the maximum possible lockdown period for a varying number of individuals within a fixed amount of resource *R*_0_. The figure reveals that with the increasing percentage of home isolation individuals, the maximum lockdown period significantly decreases for all considered cases. The reason behind this is that the more individuals are in home isolation, the more it is necessary to pay the incentives. With a fixed amount of resources, a country with less individuals can survive a longer lockdown period. With more percentages of home isolation individuals, the number of loackdown period is less, and possible of spreading of COVID-19 is also less. Therefore, the governments can consider a trade-off between increasing expenditure as a incentive and lockdown period. For 1, 000 individuals, the maximum possible lockdown period for varying amount of *R*_0_ and *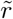* is presented in Fig. 21. The figure also illustrates that with the increasing percentages of home isolation individuals, the continuity of the lockdown period reduces for every scenarios. However, for a particular percentage of home isolation individuals where total number of individuals are fixed, a country can continue higher lockdown period who has more am amount of resources, *R*_0_. Additionally, 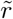 also play an important role to continue the lockdown period.

**Fig 20.**
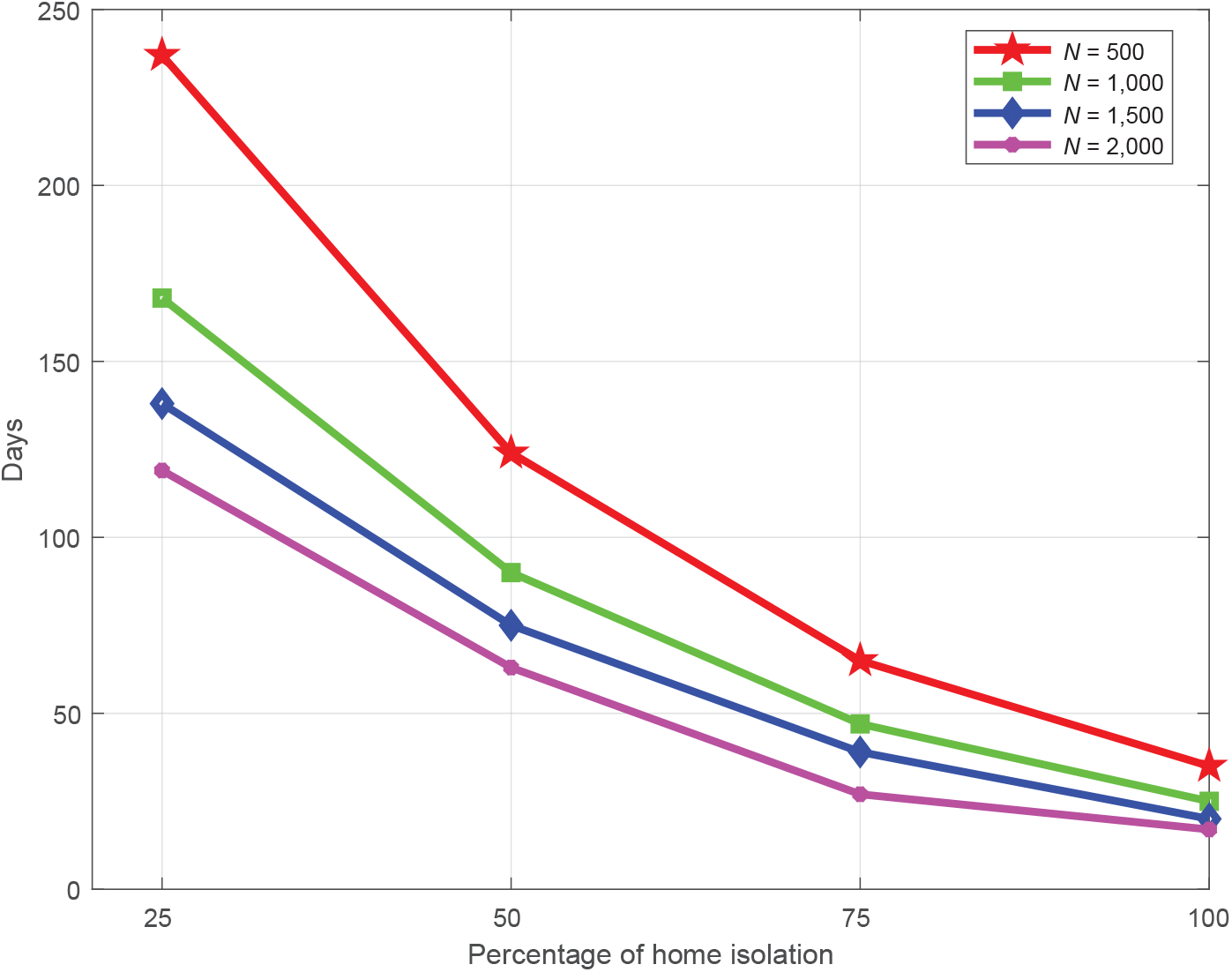
Maximum possible lockdown period with varying number of individuals when *R_0_* = 5*E* + 23, 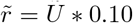, and using total incentive shown in Figure 18.

**Fig 21.**
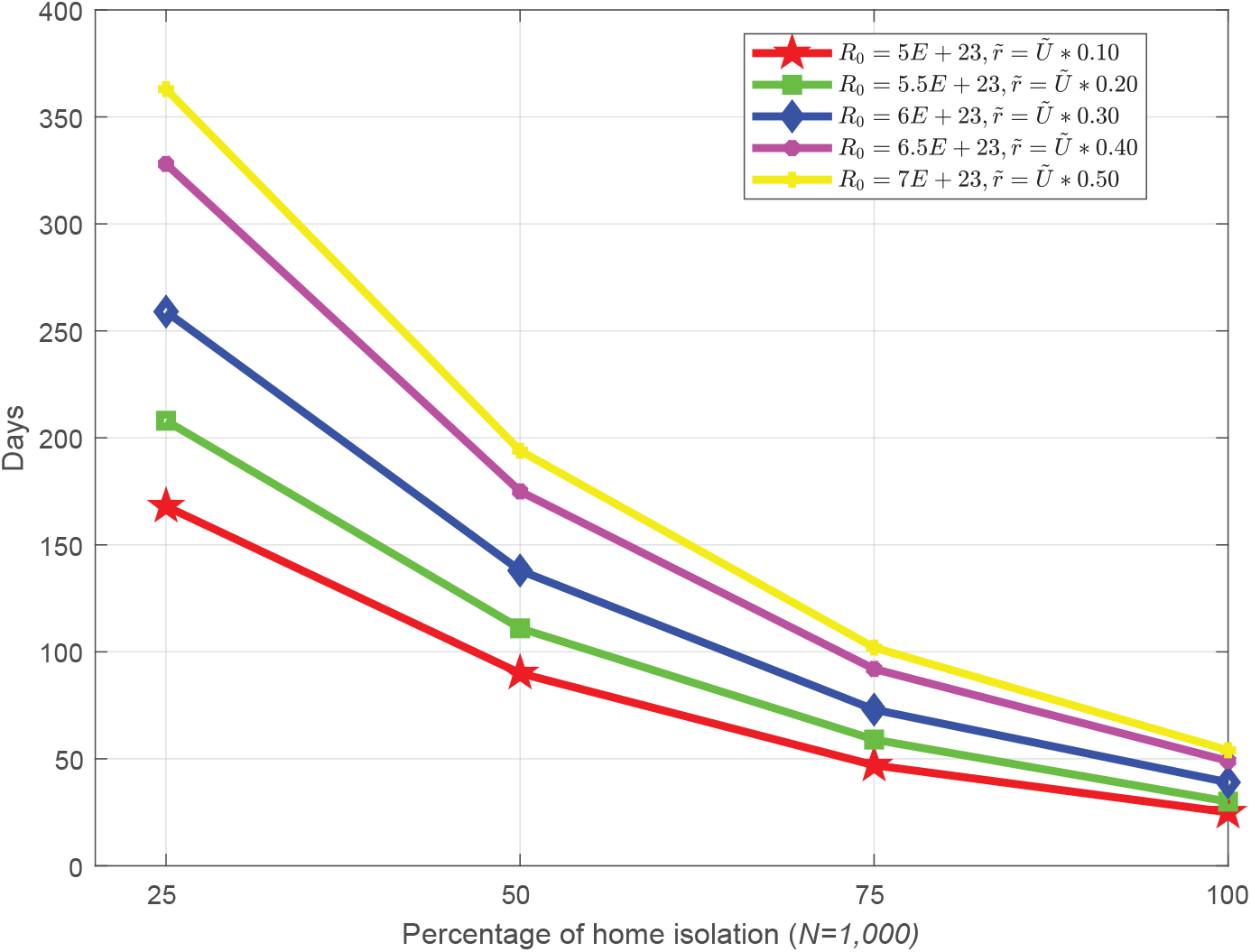
Maximum possible lockdown period with varying *R_0_* and 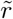 with total payoff shown in Figure 18.

## 8 Conclusions

In this paper, we have studied an analytical model for controlling the outbreak of COVID-19 by augmenting isolation and social distancing features of individuals. Here, we have observed that staying home (home isolation) is the best strategy of every individual and there is a Nash equilibrium of the noncooperative game. We have also analyzed the sustainability period of a country with a lockdown policy. Finally, we have performed a detailed numerical analysis of the proposed model to control the outbreak of the COVID-19. In future, we will further study and compare with extended cases such as centralized and different game-theoretic models. In particular, an extensive analysis between the government-controlled spread or people controlled spread under more diverse epidemic models.

## Data Availability

Wold Health Organization, “Rolling updates on event of novel coronavirus 2019 disease (covid-19),” Available Online: https://www.who.int/emergencies/diseases/novel-coronavirus-2019/ events-as-they-happen, (Accessed on 20 April, 2020).
Worldmeter, “COVID-19 Coronavirus Pandemic,” Available Online: https://www.worldometers.info/coronavirus/
Johns Hopkins University Medicine, “Coronavirus resource center,” Available Online: https://coronavirus.jhu.edu/ (Accessed on April 20, 2020).
European Centre for Disease Prevention and Control, “Situation update worldwide,” Available Online: https://www.ecdc.europa.eu/en/geographical-distribution-2019-ncov-cases, (Accessed on April 20, 2020).

https://www.who.int/emergencies/diseases/novel-coronavirus-2019/events-as-they-happen

https://www.ecdc.europa.eu/en/geographical-distribution-2019-ncov-cases

